# Assessing the impact of non-pharmaceutical interventions on SARS-CoV-2 transmission in Switzerland

**DOI:** 10.1101/2020.05.04.20090639

**Authors:** Joseph. C. Lemaitre, Javier Perez-Saez, Andrew S. Azman, Andrea Rinaldo, Jacques Fellay

## Abstract

Following the rapid dissemination of COVID-19 cases in Switzerland, large-scale non-pharmaceutical interventions (NPIs) were implemented by the cantons and the federal government between February 28 and March 20. Estimates of the impact of these interventions on SARS-CoV-2 transmission are critical for decision making in this and future outbreaks. We here aim to assess the impact of these NPIs on disease transmission by estimating changes in the basic reproduction number (R_0_) at national and cantonal levels in relation to the timing of these NPIs. We estimate the time-varying R_0_ nationally and in twelve cantons by fitting a stochastic transmission model explicitly simulating within hospital dynamics. We use individual-level data of >1,000 hospitalized patients in Switzerland and public daily reports of hospitalizations and deaths. We estimate the national R_0_ was 3.15 (95% CI: 2.13-3.76) at the start of the epidemic. Starting from around March 6, we find a strong reduction in R_0_ with a 85% median decrease (95% quantile range, QR: 83%-90%) to a value of 0.44 (95% QR: 0.27-0.65) in the period of March 29-April 5. At the cantonal-level R_0_ decreased over the course of the epidemic between 71% and 94%. We found that reductions in R_0_ were synchronous with changes in mobility patterns as estimated through smartphone activity, which started before the official implementation of NPIs. We found that most of the reduction of transmission is due to behavioural changes as opposed to natural immunity, the latter accounting for only about 3% of the total reduction in effective transmission. As Switzerland considers relaxing some of the restrictions of social mixing, current estimates of R_0_ well below one are promising. However most of inferred transmission reduction was due to behaviour change (<3% due to natural immunity buildup), with an estimated 97% (95% QR: 96.6%-97.2%) of the Swiss population still susceptible to SARS-CoV-2 as of April 24. These results warrant a cautious relaxation of social distance practices and close monitoring of changes in both the basic and effective reproduction numbers.

## Introduction

As of April 19, 2020, the ongoing coronavirus disease 2019 (COVID-19) pandemic caused by Severe Acute Respiratory Syndrome Coronavirus 2 (SARS-CoV-2) has resulted in more than 2.2 million cases and 150,000 deaths globally (WHO 2020). Independent estimates of the basic reproduction number R_0_ for SARS-CoV-2 from the initial phases of the epidemic in China, Europe and the US have generally ranged from 2-3 with doubling times on the order of 2-4 days. In reaction to rapid increase in reported cases and hospitalizations, most countries have implemented non-pharmaceutical interventions (NPIs) including compulsory face mask use, border and schools closures, quarantine of suspected and confirmed cases, up to population-wide home isolation (HIT COVID Team 2020). An observation study conducted in Hong Kong during this pandemic estimated that social distancing measures and school closures reduced COVID-19 transmission, as characterized by the effective reproductive number, by 44% (Cowling et al. 2020). Comparable reductions have been observed in many different settings (Gatto et al. 2020; Imperial College COVID-19 Response Team 2020). Making decisions around relaxing NPIs requires both a careful assessment of the level of pre-relaxation transmission (e.g., R_0_) and quantification of the expected increase in transmission from relaxation of different NPIs.

From the initial reported case on the 24th of February through April 29, Switzerland reported more than 24,400 laboratory-confirmed COVID-19 cases and 1,408 official deaths affecting all 26 cantons (OFSP 2020b). The federal government issued a series of special decrees from February 28th banning gatherings of more than 1000 people culminating on March 20 with recommended home isolation (Figure 1). One month after the first NPI, daily confirmed case incidence has decreased from a peak of more than 1000 to an average of under 170 in the week of 20-26th of April (Figure 1). Initial reports have suggested a basic reproduction number (R_0_) of 3.5 at the start of the epidemic, with a decrease of 85% by March 20 (Seth Flaxman et al. 2020). However, these estimates, part of a multi-country analysis of NPIs, relied on death incidence and did not account for specificities of the hospitalization processes in Switzerland. Moreover, changes in R_0_ were imposed to be on the date of NPI implementation, thus not allowing for the exploration of the relative timing between NPIs and changes in transmission. Furthermore, such delays might bias the estimate R_0_.

**Figure 1:**
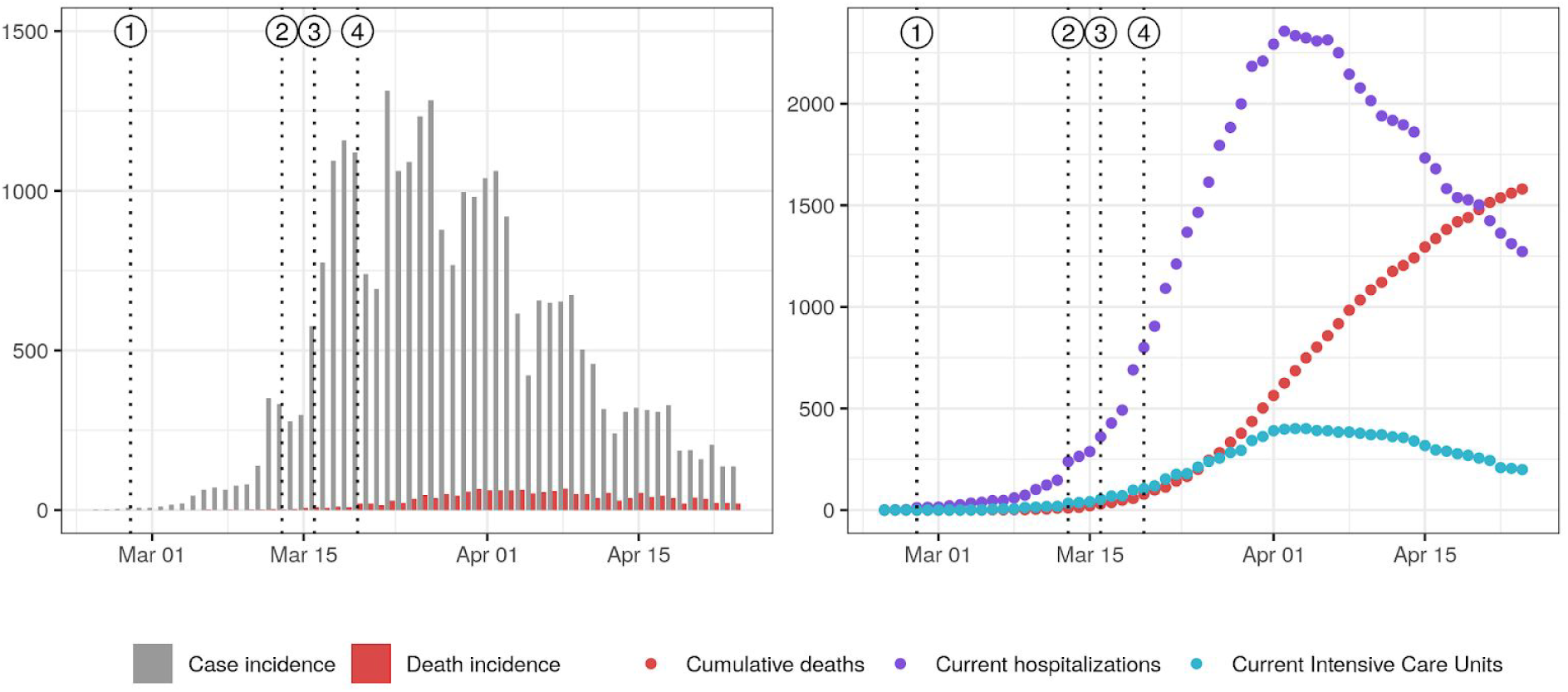
COVID-19 epidemic curve in Switzerland and timing of non-pharmaceutical interventions. Vertical dotted lines indicate the issuing of NPIs: 1) ban on gatherings of more than 1000 people, 2) school closure, 3) closure of non-essential activities, 4) ban on gatherings of more than 5 people: Left. Daily case and death incidence. Right: Current hospitalizations, Intensive Care Units (ICUs) and cumulative deaths. Plotted data was taken from (openZH 2020) and may therefore present inconsistencies with official reports from the Federal Office of Public Health due to reporting delays.

NPIs affecting daily activities such as school closures and gathering bans aim at having a direct impact on mobility patterns to reduce potentially infectious social contact. In other terms, the causal pathway from NPIs to transmission reduction is mediated by changes in mobility. Recent releases of mobility data from smartphone software providers open the possibility to study associations between the implementation of movement-limiting measures, behavioral change and the related changes in R_0_. Context-specific data on the degree and speed of compliance to these types of NPIs and associations with the observed decreases in R_0_ could inform scenario-building should the future reinstatement of measures become necessary.

Here, we aim at estimating changes in R_0_ over the course of the epidemic at both the national- and cantonal-levels using detailed data on hospitalizations and deaths from Switzerland. To begin to understand the estimated changes in R_0_, we explore its relationship with the timing of NPIs and human mobility estimates derived from cell phone data.

## Methods

### Model and assumptions

We developed a stochastic compartmental model of the COVID-19 epidemic and hospitalization processes in each canton of Switzerland. We structure our model around the classical *S, E, I* and *R* compartments (Kermack and McKendrick 1927). Namely, we consider that the population is divided in compartments depending on their status with regard to COVID-19. A susceptible individual (*S*) might be exposed after contact with infectious (*I*) individuals. Upon exposition, our former susceptible goes through an incubation period (*E*) before becoming infectious (*I*). Then he recovers (*R*) and does not participate in transmission anymore. In addition to these dynamics, in the proposed framework after individuals are infected some proportion develop severe disease and among those, some are hospitalized (H compartments) and may advance to needing the intensive care unit (U compartments). Hospitalized can progress to discharge, ICU or death and those in the ICU (U compartments) can either be discharged or die.

We only use hospitalization and death data (see below), thus we do not need to explicitly model a latent stage where individuals are still asymptomatic but infectious (Ganyani et al. 2020; He et al. 2020; Liu et al. 2020). Instead, we parameterize the model conditioning on a mean generation time of 5.2 days (Ganyani et al.), and an exposed and non-infectious duration of 2.9 days (He et al. 2020), yielding a mean duration of 4.6 days in the infectious compartments. The exhaustive description of model transitions and parameters are presented in the Supplementary Material (SM) Table 3 of the Appendix. Regarding the natural history of SARS-CoV-2, we assume that the proportion of severe infections that would require hospitalization is of 7.5%, that 50% of deaths happen outside of hospitals (data from cantons of Vaud described in section 1 of the Appendix and Geneva from OpenZH), that the hospitalized case fatality ratio is of 11% (data from canton of Vaud, SM Figure 2), and an population-level infection fatality ratio (IFR) of 0. 75 % which is in the range of published estimates (Verity et al. 2020; Russell et al. 2020).

**Figure 2:**
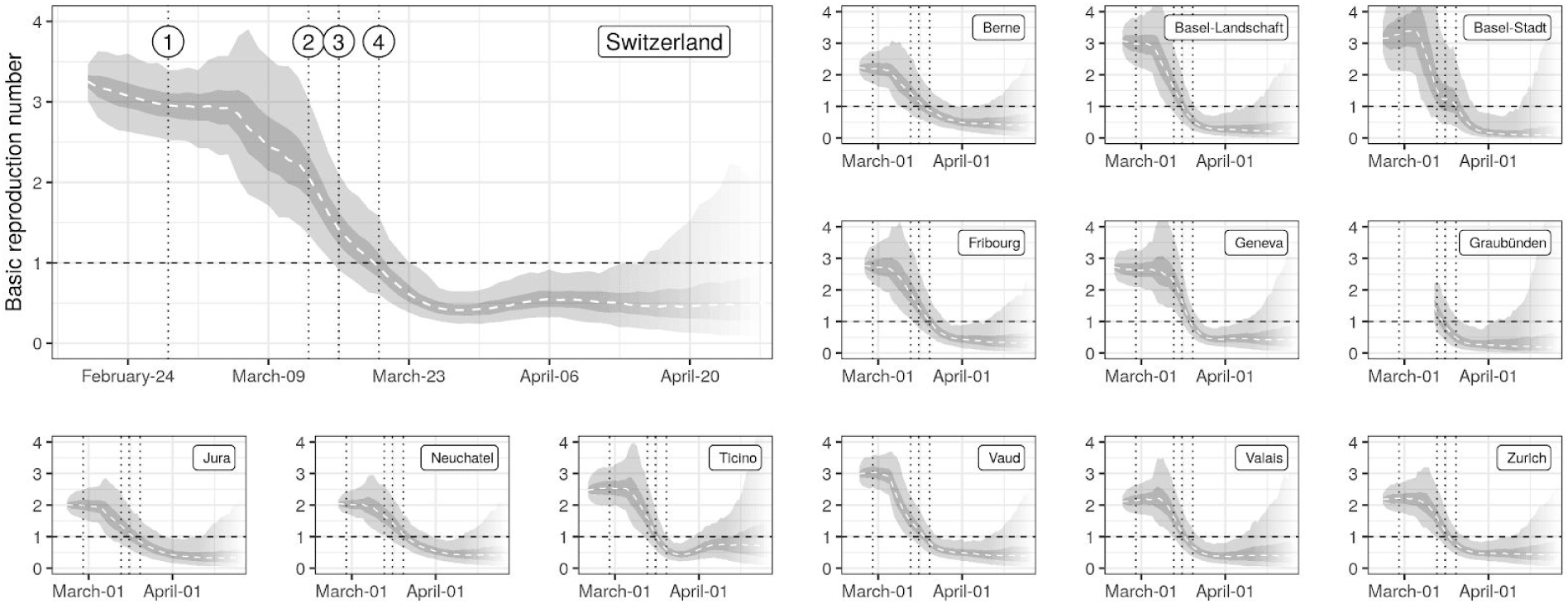
Estimates of changes in the basic reproduction number R_0_. Median (dashed line), IQR (dark gray) and the 95% QR (light gray) of the estimated time series of R_0_ are shown for each canton. Vertical dotted lines indicate the issuing of NPIs as described in Figure 1. Transparency at the end of the time series indicates increasing uncertainty (style inspired by CMMID^1^).

We used individual-level data on hospitalized cases from the canton of Vaud to estimate the distribution of time spent in the hospital and in the intensive care unit (ICU). We observed a bimodal distribution of stays in ICU and an excess of short duration of hospital stays. An Erlang mixture distribution fit to each of these durations showed support for two distinct groups with different shape and rate parameters. We therefore divided both H and U into subgroups (see section 2 of the Appendix). The number of compartments for each hospitalization state was based on distribution of timings from the Vaud data.

### Data and Inference

We use curated data from OpenZH (openZH 2020) up to April 24. This dataset includes, by canton, the number of currently hospitalized COVID patients, and the cumulative numbers of deaths, cases and hospital discharges. The latter is not available for all cantons. For our cantonal estimate, we focused on cantons which had enough cases and data to obtain meaningful results, keeping twelve of the 26 cantons (Bern, Basel-Landschaft, Basel-Stadt, Fribourg, Geneva, Graubünden, Jura, Neuchâtel, Ticino, Vaud, Valais and Zürich). These canton account for 68% of the Swiss population. Our national estimate uses national aggregate data and thus encompasses all cantons.

We fit unknown parameters of our models using maximum likelihood inference through iterated filtering (Edward L. Ionides, Bretó, and King 2006). We did not attempt to include confirmed case data into the observation model due to heterogeneous testing strategies adopted across cantons and over time. We therefore fit the model to death incidence and changes in current hospitalizations using appropriate likelihood functions (details in section 3 of the Appendix). Hospitalization incidence data at the cantonal level would have provided more information, however these data were not available at the cantonal level from OpenZH. Fitting to hospitalization incidence data, which we had access to in the canton of Vaud, yielded similar results than when using changes in current hospitalizations.

Time-varying basic reproduction numbers R_0_ were estimated following a similar approach to Cazelles et al., 2018 (Cazelles, Champagne, and Dureau 2018), recently applied to COVID-19 transmission in Hubei (Kucharski et al. 2020). The method aims at inferring the time series of R_0_ that yield model dynamics which are in best agreement with the whole set of available observations (details in section 2 of the Appendix). As such, the value of R_0_ at a given point in time is informed by the whole data, and therefore does not have the limitations of being either “forward looking” or “backwards looking” as it is the case of commonly used statistical methods applied for this purpose (Wallinga and Teunis 2004; Cori et al. 2013). Once the time series is inferred, we assess the timing and slope of changes in R_0_ by using linear changepoint models (Lindeløv 2020). The null model corresponds to a linear decrease between two plateaus corresponding to the baseline value at the start of the epidemic and a low value after the implementation of NPIs. To allow for different slopes in the decreasing phase of R_0_, we also fit models with one and two additional breakpoints (corresponding to two and three different slopes), and the best model is selected using Bayesian model selection based on leave-one-out cross-validation (details in section 6 of the Appendix).

We contrast the estimated changes in R_0_ with changes in activity-related mobility data produced by Google (Google LLC 2020). Changes in activity are expressed as relative changes with respect to a baseline computed as the median over a 5-week period from January 3 to February 6, 2020. Mobility changes were computed for different categories: grocery & pharmacy, parks, transit stations, retail & recreation, residential and workplace. Mobility estimates are based on smartphone-based geo-location location data (GPS, WiFi connections, Bluetooth) from users who activated Location History for their Google Account. These data are used to determine changes in the number of visits to and length of stays in locations categorized into the above-mentioned types. The dataset therefore only covers a sample of the Swiss population using smartphones, the latter representing around 80% of the total population in 2020 (O’Dea 2020). We use linear interpolation to fill gaps in the dataset, and we apply a 7-day moving average to smooth out weekly seasonality in activities. We compute the cross-correlation between changes in R_0_ and the averaged changes in each type of activity up to a lag of 10-days. Changes in R_0_ were computed based on location-specific baselines taken as the mean value of R_0_ from the beginning of the simulations, five days before the first reported case in each canton, until March 8th. As for R_0_ we employ changepoint models to identify dates of change in mobility patterns.

All data and code except for individual hospitalization data from Canton of Vaud have been deposited on Zenodo (doi).

## Results

Overall we find that over the study period R_0_ trends followed a common topology nationally and across cantons, starting with a high plateau (R_0_ > 2) in the early stage of the epidemic followed by a gradual, although fast, reduction starting beginning of March reaching a low and stable value (R_0_ < 1) from ends of March onwards (Figure 2).

We estimate that at the beginning of the epidemic R_0_ was 3.15 (95% CI: 2.2-3.8) at the national level, with cantonal-level values ranging from 2.0 to 3.3 (SM Table 5, excluding Graubünden for which the initial R_0_ could only be obtained on March 10th due to the absence of hospitalizations). The onset of reduction was estimated to be between March 2 (Basel Stadt and Vaud) and March 11 (Ticino) at the cantonal level and on the 6th of March at the national level (SM Figure 10). Overall we found strong support for the reduction in R_0_ starting before school closures on March 13 (probability 0.98 at the national level, SM Figure 10). Once started, we estimate a strong decrease in R_0_ at the national (reduction of 0.14/day) and cantonal levels (between 0.07/day in Jura to 0.19/day in Basel-Landschaft) (Figure 2). We did not find strong support for changes of slope during the decrease phase neither at the national nor cantonal levels except for Basel Stadt and Jura (SM Table 7). We estimate that R_0_ in Switzerland dropped below 1 on March 19 (95% CI: March 16-March 23) with individual cantons meeting this threshold between March 15 (Basel-Stadt and Grisons) and March 20 (Neuchâtel) (SM Figure 9). With respect to the timing of NPIs, we estimate the probability that R_0_ had already fallen below 1 as low when schools closed on March 13 (national: 0.04, cantons: from 0 in Geneva to 0.35 in Grisons), and high by the time gatherings of 5 people or more were banned on March 20 (national: 0.8, cantons: from 0.5 in Neuchâtel to 0.99 in Ticino) (SM Figure 8). The estimated plateau value of R_0_ after the reduction, i.e from March 29 to April 10, was of 0.4 (95% QR: 0.3-0.6) at the national level, with median values at the cantonal level ranging from 0.2 to 0.6 (SM Table 5). At the national level R_0_ was reduced by 85% (95% QR: 78%-91%), with median reductions ranging from 71% (Neuchâtel) to 94% (Basel-Stadt) at the cantonal level. A gradual reduction in R_0_ leading to values below one around the third week of March is consistent with the observed reduction of confirmed case incidence in early April, when we take into consideration the delays due to the incubation period, with median of 5.2 days (Lauer et al. 2020), and between symptom onset and reporting (Bi et al. 2020). Similarly, the inflection in the number of current hospitalizations and ICU usage in early April also supports R_0_ dropping below one in mid March.

Activity-related mobility patterns changed markedly in all cantons since the beginning of the epidemic (Figure 3). Mobility related to work, retail and recreation, and transit stations dropped by 50 to 75% at the national level, with cantonal-level reductions ranging from 30 to 80% depending on activities. Residential-related mobility increased across cantons between 20% and 30%. We find strong support for mobility changes starting simultaneously for all activity types within each canton. We estimate that changes in mobility started between March 6 and 14 for all cantons (SM Figure 11), thus finding strong support for changes starting before school closure on March 13 (national-level mean probability across activities: 0.70, cantonal range: 0.55-0.99).

Based on our changepoint models, we find that reductions in R_0_ likely started (probability: 0.76) before observed reductions in mobility at the national level and across cantons (Figure 3). Changes in R_0_ were highly correlated with changes in mobility, the strongest associations being with mobility related to work, transit stations, retail and recreation, and residential (cross-correlations > 0.9 in all cantons and nationally, Figure 4). In the majority of cases, correlation between mobility and R_0_ was strongest with no lag between the two. However, changes in mobility to workplaces lagged behind changes in R_0_ in Basel-Stadt and Basel-Landschaft. Correlations between changes in R_0_ and grocery and pharmacy mobility were less marked (national level: 0.65) with estimated lags of maximal correlation influenced by increases in travel related to groceries and pharmacies in most cantons after March 25. In Zurich and Basel-Landschaft, a strong increase in grocery and pharmacy mobility after March 25 resulted in a negative correlation with changes in R_0_ at positive lags (change in activity after change in R_0_, Figure 4). This effect is even stronger with park mobility where increases after March 25 in many cantons resulted in some cantons having negative correlations at positive lags. We did not find significant linear associations between the level of reduction in mobility and maximum reduction in R_0_ across cantons (SM Figure 7).

**Figure 3:**
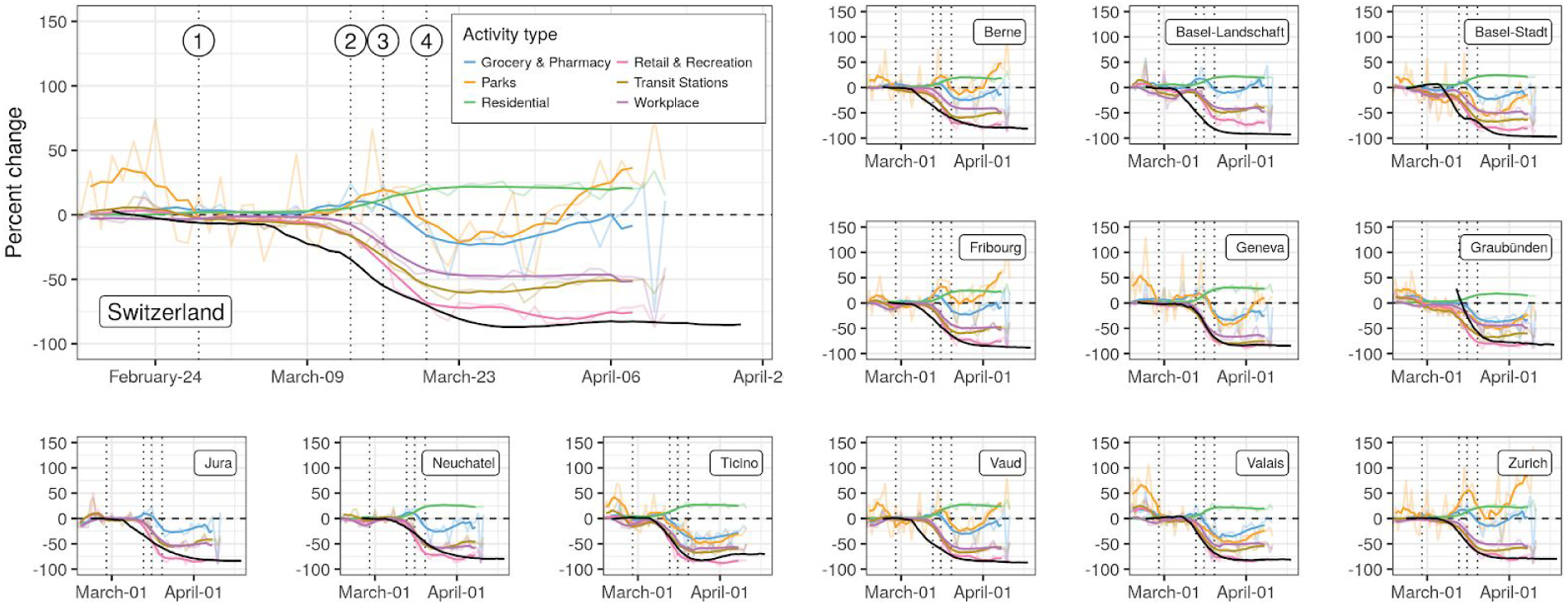
Changes in mobility patterns and R_0_. Changes in mobility with respect to baseline are shown by activity type in terms of the daily values (transparent lines) and 7-day rolling mean (full lines), against the median estimate of R_0_ (black line). Vertical dotted lines indicate the issuing of NPIs as described in Figure 1.

**Figure 4:**
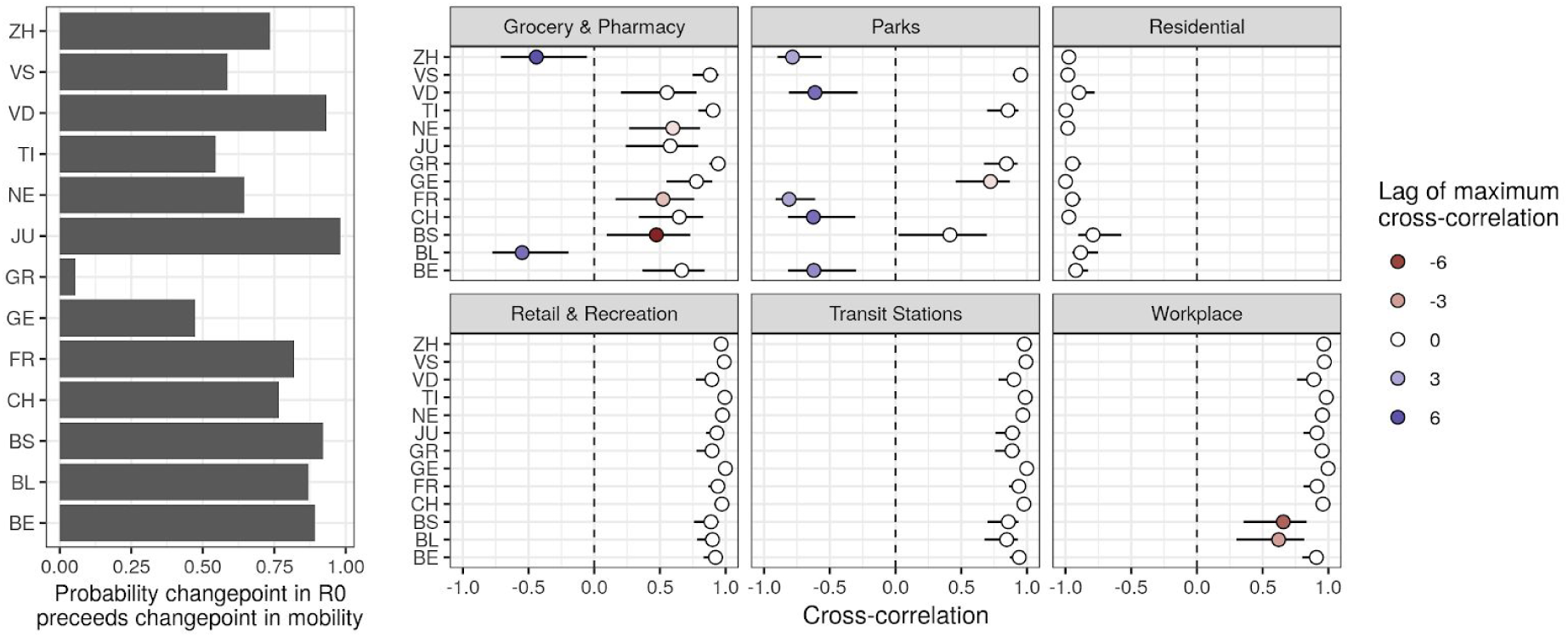
Timing between changes in R_0_ and mobility. Left: probability that the first changepoint in R_0_ occured before the first changepoint in mobility-related activity. Right: Maximum Cross-correlations between time series of changes in R_0_ and changes in mobility (bars: 95% CI). Lags refer to the delay between changes in mobility-related activity and changes in R_0_ (positive lag *k* indicates that current changes in mobility have maximal cross-correlation with changes in R_0_*k* days in the past).

We estimated a commonly reported metric, the effective reproduction number (R_eff_), which is an aggregate measure of transmission capturing aspects of both infectious contacts and of population susceptibility. Across cantons, R_eff_ was extremely close to R_0_ indicating that a small fraction of the population is expected to have natural immunity to SARS-CoV-2 (as we assumed was true in the short-term after infection). We estimate that as of April 24 3% (95% quantile range: 2.8%-3.4%) of the population nationally has been infected, with median estimates ranging from 1.5% (Bern) to 14% (Ticino) (Figure 5).

**Figure 5:**
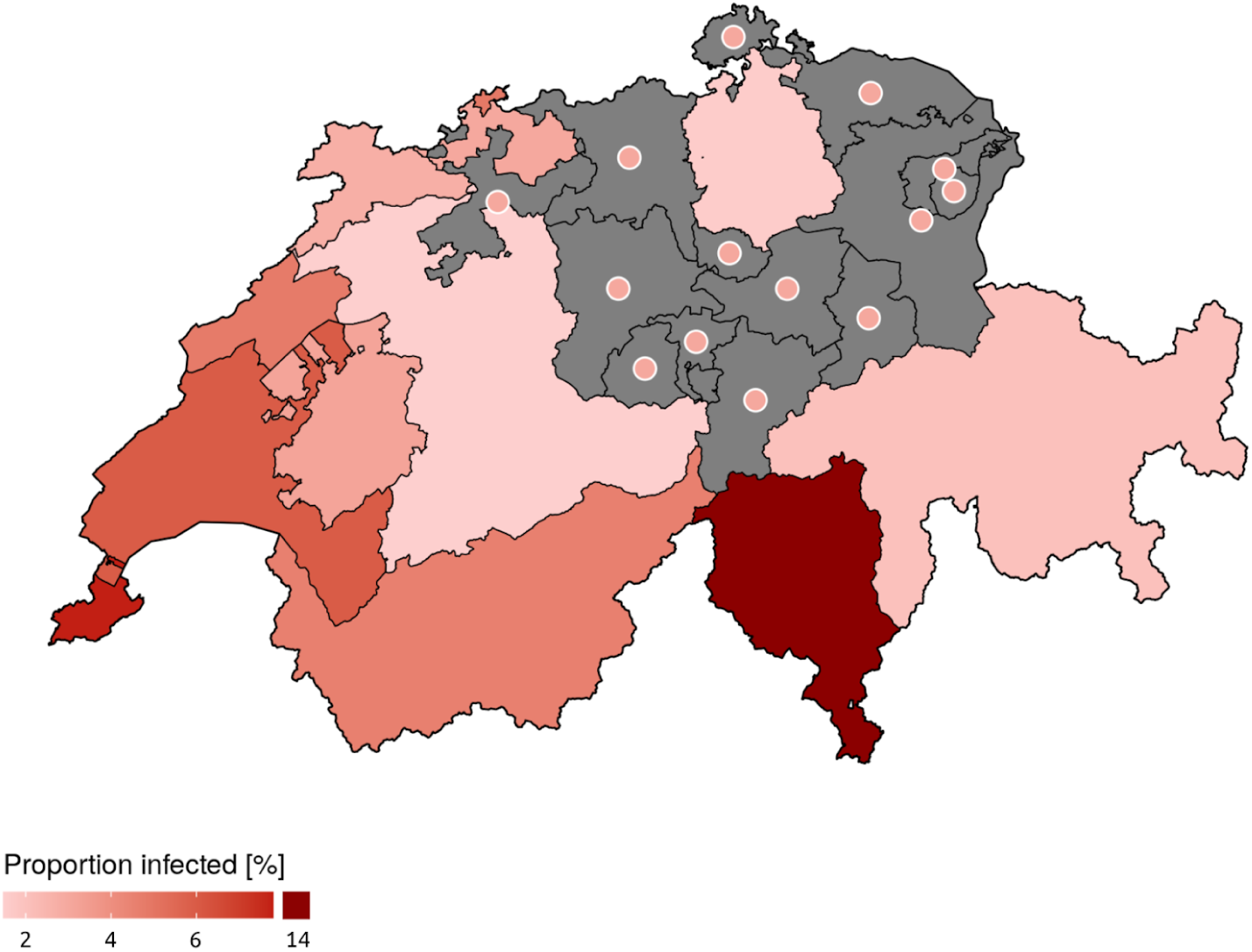
Modelled proportion of people infected with SARS-CoV-2 in Switzerland up to April 24th. Estimates were produced for 12 of 26 cantons for which enough data were available (unmodelled cantons are shown in gray with points indicating the national-level estimated incidence proportion of 3%). Values are reported in SM Table 6 of the Appendix.

## Discussion

Our results suggest a strong reduction of R_0_ across Switzerland since the start of the epidemic. The reduction in R_0_ started around March 6, thus at least one week before the implementation of lockdown-type NPIs. Analysis of activity-related mobility data also showed strong support for changes in mobility starting prior to the implementation of most NPIs. Estimated reductions of viral transmission were strongly correlated and mostly synchronous with observed changes in mobility patterns, although the initiation of changes in transmission preceded measurable changes in activity-related mobility.

The methods used to infer the time series of R_0_ do not rely on assumptions on the shape of how it changed in time, nor on the dates at which change started. Alternative methods that rely on fixed dates (such as Imperial College COVID-19 Response Team 2020) might be biased as changes in transmission are not synchronous with policy changes. Distribution based methods such as provided by R package EpiEstim (Wallinga and Teunis 2004; Cori et al. 2013) are flexible but subject to bias when misused (Lipsitch, Joshi, and Cobey [2020] 2020). In addition our approach enables the estimation of R_0_ which is a direct quantification of transmission potential, as opposed to the effective reproduction number R_eff_ which also accounts for the effect of susceptible depletion as done in the above-mentioned statistical approaches. This enables us to estimate the proportion of reduction in transmission attribuable to behavioral changes, which is therefore more suited to study the impact of NPIs. Aside from these methodological differences, our estimates are in line with other estimates in Switzerland: Althaus et al. estimate a reduction of 89 % (83% - 94%) from a baseline of 2.78 (2.51 - 3.11) (Althaus 2020), Scire et al. estimate a reduction of 76 % (70% - 82%) from a baseline of 1.88 (1.80 - 1.98) (Scire et al. 2020) and Imperial College estimates a reduction of 60% (50% - 80%) from a baseline of 3.5 (2.8 - 4.3) (Seth Flaxman et al. 2020).

Our results provide strong support for a reduction of transmission starting about 1 week prior to school closure, the first national-level NPI targeting daily activities, which was ordered on the 13th of March. Moreover, initiation of transmission reduction was found to precede changes in mobility patterns as detectable from the Google dataset. A possible explanation for this initial decrease in transmission could be linked to the strong increase in public interest in COVID-19 in late February as measured by Google searches for COVID-19-related keywords (SM Figure 13). In fact, we estimated that the sharp rise in Google searches started on March 7 (95% CrI: March 3-March 9) which overlaps with our estimated start of national level decrease in R0 on March 6 (probability that changepoints coincide: 0.64). The Federal Office for Public Health issued an information campaign on COVID-19 on February 28th which was updated on March 2 stressing basic hygiene rules (OFSP 2020a). This may have resulted in voluntary social distancing as well as increased hygiene early on without noticeable changes in mobility patterns. This type of proactive changes in behavior would be in line with early changes in mobility patterns, which were estimated to precede school closures and subsequent measures.

Our results suggest that the value of R_0_ was likely already below one on March 20 when the federal government banned gatherings of more than 5 people and recommended voluntary home isolation for the whole population. This result should however be taken within context as the announcement was anticipated on social networks earlier that week, thus probably already impacting social distancing behavior. We therefore warrant caution in any causal interpretation of our results on the role of this last NPI on driving R_0_ below one.

Despite the strong association between the changes in mobility and reductions in R_0_ within each canton, the lack of cross-cantonal associations between the level of reduction in mobility types and the level of reduction in R_0_ suggests context-specific pathways between COVID-19 transmission and mobility intensity. This warrants caution in attempting to apply general relations between mobility and transmission reduction. Investigating general associations will require more in-depth studies controlling for other factors such as population density, economic activities and social contact matrice, inter-cantonal mobility patterns, in addition to incorporating potential environmental drivers of transmission such as temperature and relative humidity (Neher et al. 2020; Kissler et al. 2020).

We note several limitations to this work. First, due to the relatively recent introduction of SARS-CoV-2 in Switzerland compared to the length of hospital and ICU stays, the time distribution of in- and out-of hospital patients is biased towards shorter duration (SM Figure 3). In addition, due to the limited data available in some places, we were only able to fit our model for twelve of the twenty-six cantons. Modeling results presented in this work are subject to our hypothesis on yet uncertain parameters of COVID-19 including the infection fatality rate and the proportion of severe infections requiring hospitalization. Ongoing serological studies in Switzerland will provide key data to narrow these uncertainties. Moreover our estimates of time-varying basic reproduction numbers assume that the generation interval for COVID-19 in Switzerland remained unchanged, thus potentially ignoring the joint role of R_0_, the infectious period and contact rates in determining the disease’s intrinsic growth rate (Yan 2008). Assuming that the generation interval increased with the reduction of social contact, our estimates are conservative over-estimates of the “true” value of R_0_, which is encouraging from a public health perspective. Another limitation of our study is that it was not possible to disentangle the individual contribution of each NPI on R_0_ in this analysis due to the early onset of changes in R_0_ and in mobility patterns as well as the very close spacing between the different types of NPIs. This information would however be extremely valuable in supporting decisions on NPI strategies against COVID-19. Efforts to constitute a global database of NPIs will open the opportunity to extend this type of analysis to other settings and produce evidence for the effect of different types of NPIs (HIT COVID Team 2020).

As the Swiss government plans to gradually lift restrictions, close monitoring of changes in R_0_ is critical given that the reductions in transmission appear to be almost entirely driven by changes in behavior, not through herd immunity. Near real-time estimates of R_0_ may serve as a critical tool for public health and political decision makers in the months to come and efforts should be made to refine models like ours using new data, including those from population-based serologic studies, mobility data and more detailed individual-level data on COVID-19 cases across the spectrum of severity

## Data Availability

All data and code except for individual hospitalization data from Canton of Vaud will be made available on Zenodo upon peer-review.

## Statement on funding sources and conflicts of interest

The authors declare no conflict of interest. JL acknowledges funds provided by the Swiss National Science Foundation via the project *“Optimal control of intervention strategies for waterborne disease epidemics”* (200021-172578). AR acknowledges funding from Fondazione Cassa di Risparmio di Padova e Rovigo (IT) through its grant 55722 (April 2020).

## Acknowledgments

We thank Canton of Vaud.

## Footnotes

1 https://cmmid.github.io/topics/covid19/global-time-varying-transmission.html [Accessed April 30th 2020]

## Notes

### Competing Interest Statement

The authors have declared no competing interest.

